# COVID-19 :Determinants of Hospitalization, ICU and Death among 20,293 reported cases in Portugal

**DOI:** 10.1101/2020.05.29.20115824

**Authors:** Vasco Ricoca Peixoto, André Vieira, Pedro Aguiar, Paulo Sousa, Carlos Carvalho, Daniel Thomas, Alexandre Abrantes, Carla Nunes

## Abstract

**Introduction:** Determinants of hospitalization, intensive care unit (ICU) admission and death are still unclear for Covid-19 and only a few studies have adjusted for confounding for different clinical outcomes including all reported cases in a country. We used routine surveillance data from Portugal to identify risk factors for COVID-19 outcomes, in order to support risk stratification, clinical and public health interventions, and scenarios to plan health care resources.

**Methods:** We conducted a retrospective cohort study including 20,293 laboratory confirmed cases of COVID-19 in Portugal extracted in April 28 2020, electronically through the National Epidemic Surveillance System of the Directorate-General of Health(DGS). We calculated absolute risks, relative risks (RR) and adjusted relative risks (aRR) to identify demographic and clinical factors associated with hospitalization, admission to ICU and death using Poisson regressions.

**Results:** Increasing age after 60 years was the greatest determinant for all outcomes. Assuming 0–50 years as reference, being aged 80–89 years was the strongest determinant of hospital admission (aRR-5.7), 70–79 years for ICU(aRR-10.4) and > 90 years for death(aRR-226.8) with an aRR of 112.7 in those 70–79. Among comorbidities, Immunodeficiency, cardiac disease, kidney disease, and neurologic disease were independent risk factors for hospitalization (aRR 1.83,1.79,1.56, 1.82), for ICU these were cardiac, Immunodeficiency, kidney and lung disease (aRR 4.33, 2.76, 2.43, 2.04), and for death they were kidney, cardiac and chronic neurological disease (aRR: 2.9, 2.6, 2.0) Male gender was a risk factor for all outcomes. There were small statistically significant differences for the 3 outcomes between regions.

**Discussion and Conclusions:** Older age stands out as the strongest risk factor for all outcomes specially for death as absolute is risk was small for those younger than 50. These findings have implications in terms of risk stratified public health measures that should prioritize protecting older people although preventive behavior is needed in all ages. Epidemiologic scenarios and clinical guidelines may consider these estimated risks, even though under-ascertainment of mild and asymptomatic cases should be considered.

## Introduction

Previous studies of clinical outcomes of COVID-19 in China^1^, Italy^2 3^ and the United States of America^4^ have described a number of factors associated with poorer clinical outcomes, including age, sex and comorbidities not adjust for confounding. Identifying these determinants and their specific risk can help inform policy on risk stratification, implementation of public health measures and improve epidemiological scenarios on the needed healthcare resources.

Based on initial data, the mean case fatality rate for adults aged under 60 is estimated to be less than 0.2%, compared with 9.3% in those aged over 80. Even if comorbidities significantly increased mortality risk significantly, risk would remain lower for younger people than for most older adults.^5^

Other studies on risk factors for clinical outcomes of COVID-19 included small series of patients, mostly among those hospitalized or excluding mild disease,^6 7 8^ making it difficult to produce reliable estimates for specific risk factors in the general population.

There is still uncertainty about the contribution of each factor for hospitalization, ICU admission and death in the general population as only a few studies conducted multivariable analysis to account for confounding^9 10 11^ One study of laboratory-confirmed Covid-19 cases in New York City found increased risks of hospitalization for those aged ≥ 75 years and aged 65–74 when compared to those 19 to 44 (OR 66.8, 95% CI, 44.7–102.6 and OR 10.9, 95% Cl, 8.3514.34, respectively), BMI>40 (OR 6.2, 95% CI, 4.2–9.3), and heart failure (OR 4.3 95% CI, 1.9− 11.2)^9^ A large UK cross-sectional survey describing 16,749 patients already hospitalized with COVID-19 showed higher risk of death for patients with increased age, cardiac, pulmonary and kidney disease, as well as malignancy, dementia and obesity^10^. The largest cohort study to date was conducted in the UK, the OpenSAFELY Collaborative study^11^, including 17 million adult NHS patients. Using a multivariable analysis, it was found that being in older age groups, being male, older and living in a more socio-economically deprived community; having uncontrolled diabetes, severe asthma and various other prior medical conditions were the most relevant risk factors for death by COVID-19. As in other studies hypertension was not a risk factor after adjustment^12 13^.

High quality data on possible risk factors for poor outcomes of COVID-19 is needed to inform public health policy and preparedness. Portugal may be well placed to provide such data and risk estimates as high cases ascertainment for mild cases may improve risk measures for outcomes in the general population. It is a country with a relatively high case ascertainment estimates, (from 22% (18% – 39%)^14^ to 36.6% (29.5% – 45.7%)^15^) and had one of higher testing rates in mid-May^16^ despite having lower transmission levels than many countries^17^. Adding to this since there is a National Surveillance System^18^ implemented since 2015, we believe this study may contribute to further understanding population level COVID-19 risk factors for three different outcomes: Hospitalization, ICU admission and death.

We aim to identify risk factors for more severe COVID-19 clinical outcomes in Portugal order to better support risk stratification, clinical and public health interventions and to plan health care resources.

## Methods

### Study design

A retrospective cohort study including all reported cases of COVID-19 in Portugal was conducted. Outcomes were: Hospitalization, ICU admission and death. We calculated absolute risk, relative risk and adjusted relative risk for age, gender, comorbidities and region of residence using a Poisson Regression.

### Data sources

We obtained anonymized data from the Directorate-General of Health (DGS), including all confirmed cases of COVID-19 notified to the National Epidemiological Surveillance System (SINAVE) extracted in April 28 2020. SINAVEmed is an electronic platform in which clinicians are obliged by law to notify all suspected and confirmed cases of COVID-19, and includes questions about clinical findings and preexisting conditions. Notifications trigger an epidemiological investigation by the Local Public Health Services, where a Public Health Physician (health authority in the area of residence of the case) validates the case. At a later stage the Regional Public Health Department and finally DGS conducts a final validation of case information. Outcome data are completed primarily at the local level, but can updated at the Regional and National level(DGS).

### Case definitions

A confirmed case of Covid-19 is defined as anyone with positive result for SARS-CoV-2 RNA (by RT-PCR) in nasopharyngeal and/or oropharyngeal specimens.

To March 26, all patients with fever or cough and contact with a symptomatic case or returning from an active transmission zone (outside Portugal) were considered suspect cases and had indication to be tested. From March 26, all people with fever or cough regardless of epidemic link were considered suspect and told by public health officials to call the NHS Health line and were subsequently sent for testing.

### Outcomes

We evaluated three primary outcomes: hospitalization in general ward(not in ICU), admission to intensive care Unit (ICU) and death. Outcomes were considered according to data from SINAVE filled as described above according to patients situation and clinical evolution.

### Risk Factors

From SINAVEmed dataset we included the following variables: age, sex, chronic diseases/ comorbidities (Asthma, Cancer, Cardiac Disease, Diabetes, Immunodeficiencies(including HIV), Kidney Disease, Liver Disease, Lung Disease(other than Asthma), Hematologic Disease, Chronic Neurologic Disease(including dementia) and region of occurrence of the case. Regions were included for adjustment. Norte, Centro Lisboa e Vale do Tejo (LVT), Algarve, Alentejo, Açores, Madeira.

### Statistical analysis

Descriptive statistics were applied to characterize the cohort of confirmed COVID-19 cases and the distribution by outcomes. We conducted univariable analysis and calculated absolute risks (proportion were each outcome was observed by stratum), relative risk (RR) with95% confidence intervals (CI) and p-value (Wald).We then calculated fully adjusted relative risk (aRR) through multivariate analysis, using Poisson regression models that included the same co-variables for each outcome.

Age was categorized in 6 categories with reference from 0–50 and then 10 years age groups until > 90

Finally we built forest plots with aRR for the Poisson Regression and CI for the 3 clinical outcomes analyzed.

To make explicit the assumptions behind variables included in the models we drew a Directed Acyclic Graph on the relations between variables and potentially biasing pathways.(Figure 1.)

**Figure 1.**
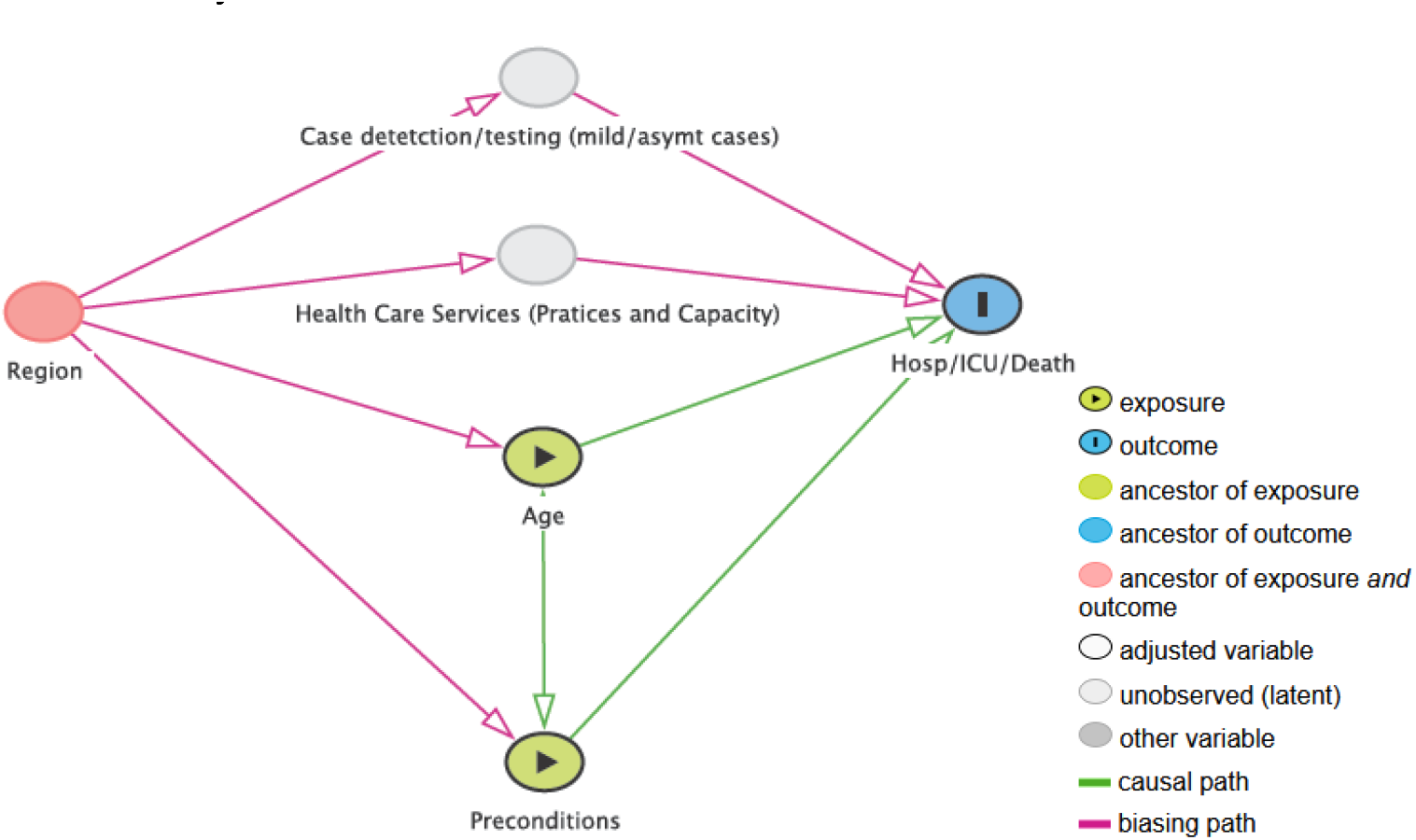
Directed Acyclic Graph representing the assumptions in relationships between variables for adjustment.

**Figure 2.**
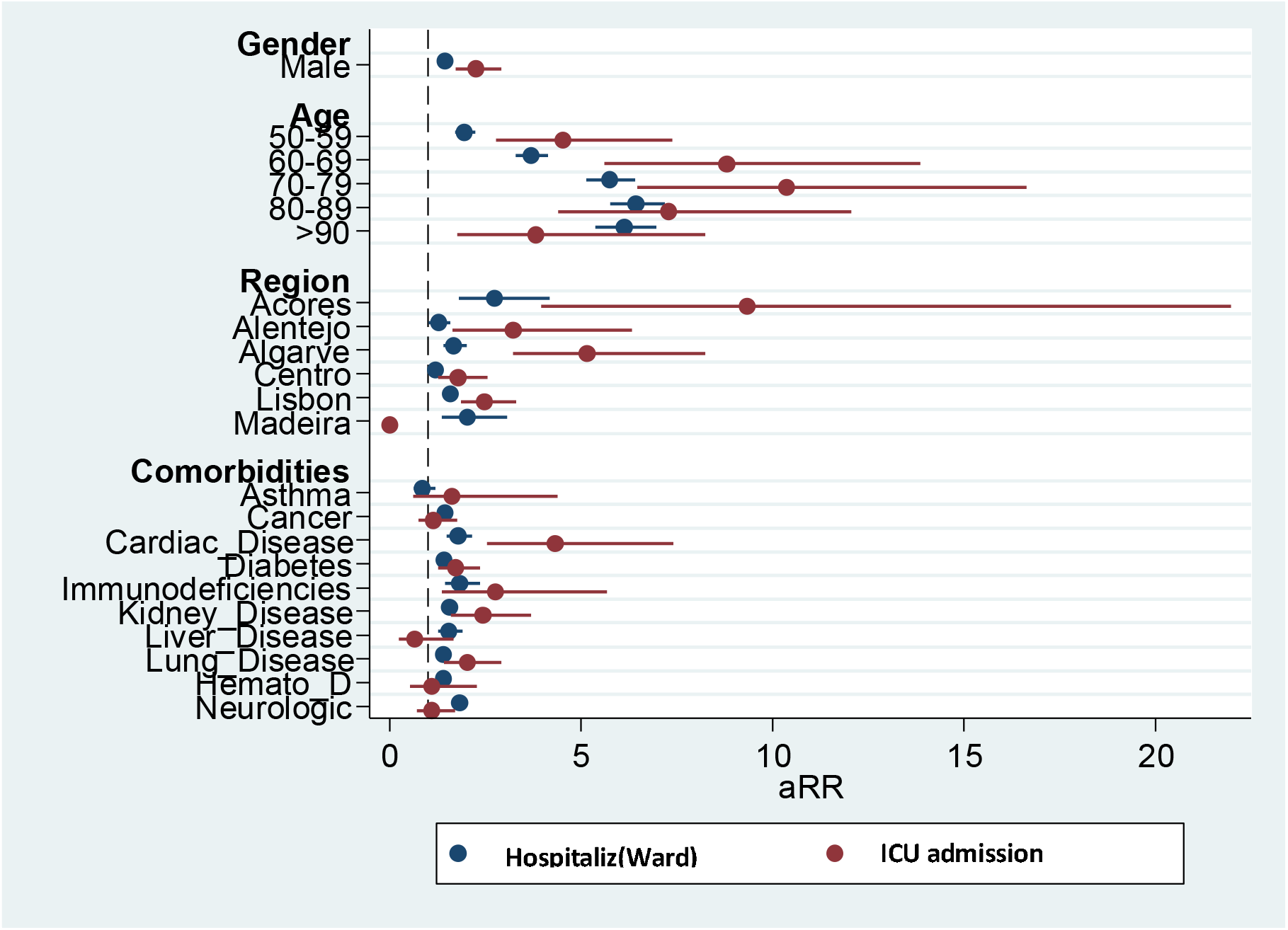
Forest Plot for Poisson aRR for hospital admission and ICU admission (Reference categories: Gender(Female); Age(0−49);Region: North)

**Figure 3.**
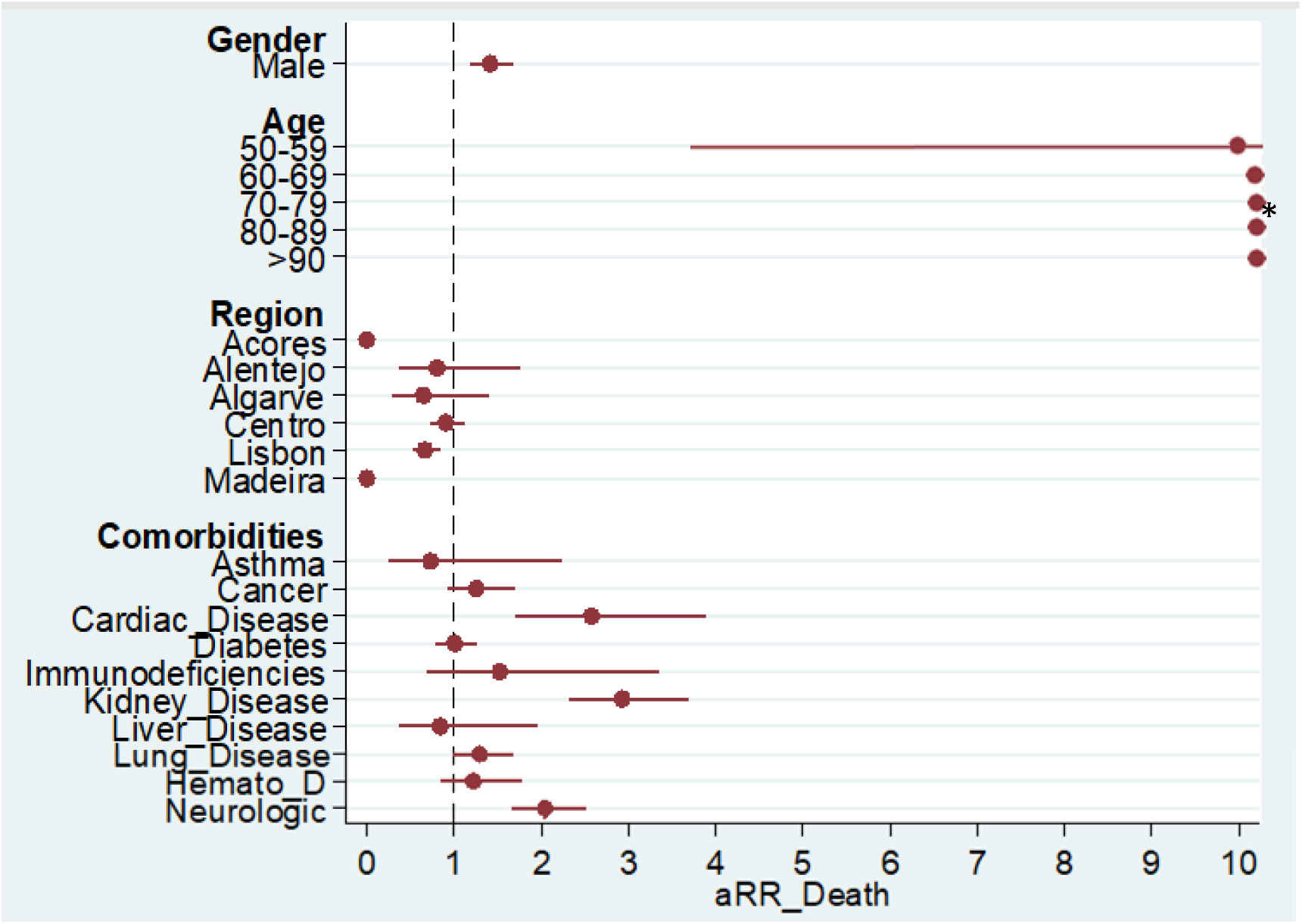
Forest Plot for Poisson aRR for Death ( Age Risk after 50-59 in footnote; Reference categories: Gender(Female); Age(0−49);Region: North)

The regression models were analyzed in STATA 14, All analyses used 95% CI and < 0.05 as statistically significant.

### Ethical Considerations

Data was shared by DGS with the National School of Public Health under a partnership for COVID-19 research. Ethical Committee of the National School of Public Health approved the project (Approval: CE/ENSP/CREE/2/2020).

## Results

Among 20,293 laboratory confirmed cases of COVID-19,2972 (14.6%) were admitted to hospital(general ward), 261 (1.3%) were admitted to an ICU and 502 (2.5%) died. Among cases 42% were male, 52% of cases were above 50 years and 17% had at least one comorbidity recorded. Northern Region had 60% of cases.

### Risk Factors for Hospitalization

In the univariable analysis Absolute risk of Hospitalization increases with age and it is the strongest risk factor. Age groups above 60 presents aRR of hospitalization higher than any chronic disease alone after adjustment assuming 0–50 reference. For Age 70–79 aRR was 5.737 (CI95%:5.133–6.412)

Different regions had different hospitalization risks and statistically significant differences in RR that are maintained after full adjustment. Preexisting conditions with higher adjusted RR were Immunodeficiencies (aRR:1.831 CI95%: 1.434–2.346), Cardiac Disease (aRR:1.79 CI95%:1.489− 2.152), Kidney Disease (aRR:1.557 CI95%:1.415–1.713), Liver Disease (aRR:1.543 CI95%: 1.247−1.91) and Neurologic Disease aRR(1.82 CI95%1.69–1.959). (Table 1)

**Table 1.** Association between geodemographic factors and pre-existing conditions and hospitalization (general ward)

| Exposure | Total | Hosp | Hosp% | Crude RR | CI95% | P-value | aRR | CI95% | p-value |
| --- | --- | --- | --- | --- | --- | --- | --- | --- | --- |
| **Sex** |  |  |  |  |  |  |  |  |  |
| Female | 10949 | 1416 | 12.93 |  |  |  |  |  | Ref |
| Male | 7721 | 1556 | 20.15 | 1.56 | [1.46-1.66] | <0.001 | 1.431 | [1.346-1.521] | <0.001 |
| **Age Group (Ref-0-9)** |  |  |  |  |  |  |  |  |  |
| 0-49 | 9055 | 462 | 5.1 |  |  |  |  |  | Ref |
| 50-59 | 3325 | 336 | 10.11 | 1.98 | [1.73-2.27] | <0.001 | 1.948 | [1.707-2.222] | <0.001 |
| 60-69 | 2233 | 491 | 21.99 | 4.31 | [3.83-4.85] | <0.001 | 3.691 | [3.284-4.148] | <0.001 |
| 70-79 | 1653 | 659 | 39.87 | 7.81 | [7.02-8.69] | <0.001 | 5.737 | [5.133-6.412] | <0.001 |
| 80-89 | 1678 | 747 | 44.52 | 8.73 | [7.87-9.68] | <0.001 | 6.437 | [5.766-7.185] | <0.001 |
| >90 | 726 | 277 | 38.15 | 7.48 | [6.58-8.50] | <0.001 | 6.12 | [5.379-6.963] | <0.001 |
| **Region (Ref- North)** |  |  |  |  |  |  |  |  |  |
| North | 11090 | 1453 | 13.1 |  |  |  |  |  | Ref |
| Acores | 48 | 14 | 29.17 | 2.23 | [1.43-3.47] | 0.001 | 2.742 | [1.804-4.169] | <0.001 |
| Alentejo | 370 | 54 | 14.59 | 1.11 | [0.87-1.43] | 0.403 | 1.269 | [1.019-1.58] | 0.033 |
| Algarve | 462 | 92 | 19.91 | 1.52 | [1.26-1.84] | <0.001 | 1.672 | [1.393-2.007] | <0.001 |
| Center | 2651 | 510 | 19.24 | 1.47 | [1.34-1.61] | <0.001 | 1.19 | [1.094-1.293] | <0.001 |
| LVT | 3951 | 827 | 20.93 | 1.6 | [1.48-1.73] | <0.001 | 1.582 | [1.477-1.695] | <0.001 |
| Madeira | 87 | 15 | 17.24 | 1.32 | [0.83-2.09] | 0.255 | 2.034 | [1.353-3.056] | 0.001 |
| **Comorbidities** |  |  |  |  |  |  |  |  |  |
| Asthma | 258 | 26 | 10.08 | 0.63 | [0.44-0.91] | 0.01 | 0.859 | [0.622-1.186] | 0.356 |
| Cancer | 579 | 292 | 50.43 | 3.4 | [3.12-3.72] | <0.001 | 1.44 | [1.308-1.585] | <0.001 |
| Cardiac_Disease | 52 | 49 | 94.23 | 6 | [5.57-6.47] | <0.001 | 1.79 | [1.489-2.152] | <0.001 |
| Diabetes | 1057 | 496 | 46.93 | 3.34 | [3.10-3.59] | <0.001 | 1.428 | [1.325-1.538] | <0.001 |
| Immunodeficiencies | 99 | 43 | 43.43 | 2.75 | [2.19-3.46] | <0.001 | 1.834 | [1.434-2.346] | <0.001 |
| Kidney_Disease | 382 | 273 | 71.47 | 4.84 | [4.50-5.21] | <0.001 | 1.557 | [1.415-1.713] | <0.001 |
| Liver_Diseas | 102 | 64 | 62.75 | 4.01 | [3.44-4.67] | <0.001 | 1.543 | [1.247-1.91] | <0.001 |
| Lung_Disease | 637 | 292 | 45.84 | 3.08 | [2.82-3.38] | <0.001 | 1.399 | [1.273-1.537] | <0.001 |
| Hemato_Disease | 202 | 131 | 64.85 | 4.22 | [3.79-4.69] | <0.001 | 1.4 | [1.226-1.599] | <0.001 |
| Neurologic_Disease | 733 | 476 | 64.94 | 4.67 | [4.38-4.98] | <0.001 | 1.82 | [1.69-1.959] | <0.001 |

**Table 2.** Association between geodemographic and pre-existing conditions and admission to Intensive Care Unit

| Exposure | Total | ICU | ICU(% within stratum) | crudeRR | CI95% | P-value | aRR | CI95% | p-value |
| --- | --- | --- | --- | --- | --- | --- | --- | --- | --- |
| **Gender** |  |  |  |  |  |  |  |  |  |
| Female | 11903 | 88 | 0.74 |  |  |  |  |  | Ref |
| Male | 8390 | 173 | 2.06 | 2.79 | [2.16-3.60] | <0.001 | 2.245 | [1.723-2.925] | <0.001 |
| **Age** |  |  |  |  |  |  |  |  |  |
| 0-49 | 9675 | 25 | 0.26 |  |  |  |  |  | Ref |
| 50-59 | 3549 | 40 | 1.13 | 4.36 | [2.65-7.18] | 0.005 | 4.532 | [2.778-7.393] | <0.001 |
| 60-69 | 2463 | 67 | 2.72 | 10.53 | [6.66-16.63] | <0.001 | 8.805 | [5.594-13.861] | <0.001 |
| 70-79 | 1808 | 70 | 3.87 | 14.98 | [9.52-23.59] | <0.001 | 10.376 | [6.47-16.641] | <0.001 |
| 80-89 | 1932 | 50 | 2.59 | 10.02 | [6.21-16.15] | <0.001 | 7.281 | [4.396-12.058] | <0.001 |
| <90 | 866 | 9 | 1.04 | 4.02 | [1.88-8.59] | 0.01 | 3.815 | [1.765-8.244] | 0.001 |
| **Region** |  |  |  |  |  |  |  |  |  |
| North | 12207 | 101 | 0.83 |  |  |  |  |  | Ref |
| Acores | 48 | 3 | 6.25 | 7.55 | [2.48-22.98] | <0.001 | 9.333 | [3.965-21.97] | <0.001 |
| Alentejo | 387 | 9 | 2.33 | 2.81 | [1.43-5.52] | 0.002 | 3.221 | [1.639-6.333] | 0.001 |
| Algarve | 472 | 18 | 3.81 | 4.61 | [2.82-7.55] | <0.001 | 5.159 | [3.23-8.242] | <0.001 |
| Center | 2812 | 44 | 1.56 | 1.89 | [1.33-2.69] | <0.001 | 1.792 | [1.259-2.549] | 0.001 |
| LVT | 4264 | 85 | 1.99 | 2.41 | [1.81-3.21] | <0.001 | 2.48 | [1.866-3.297] | <0.001 |
| Madeira | 90 | 0 | 0 | 0 | [.-.] | 0.386 | 0 | [0-0] | <0.001 |

| Comorbidities - |  |  |  |  |  |  |  |  |  |
| --- | --- | --- | --- | --- | --- | --- | --- | --- | --- |
| Asthma | 277 | 4 | 1.44 | 1.12 | [0.42-3.00] | 0.814 | 1.628 | [0.605-4.381] | 0.334 |
| Cancer | 611 | 22 | 3.6 | 2.97 | [1.93-4.55] | <0.001 | 1.143 | [0.739-1.767] | 0.549 |
| Cardiac_Disease | 54 | 10 | 18.52 | 14.93 | [8.42-26.48] | <0.001 | 4.33 | [2.53-7.411] | <0.001 |
| Diabetes | 1145 | 53 | 4.63 | 4.26 | [3.17-5.73] | <0.001 | 1.715 | [1.256-2.343] | 0.001 |
| Immunodeficiencies | 107 | 7 | 6.54 | 5.2 | [2.51-10.75] | <0.001 | 2.767 | [1.348-5.683] | 0.006 |
| Kidney_Disease | 400 | 34 | 8.5 | 7.45 | [5.27-10.53] | <0.001 | 2.439 | [1.61-3.697] | <0.001 |
| Liver_Disease | 107 | 5 | 4.67 | 3.68 | [1.55-8.75] | 0.002 | 0.643 | [0.25-1.657] | 0.361 |
| Lung_Disease | 688 | 37 | 5.38 | 4.71 | [3.35-6.61] | <0.001 | 2.035 | [1.421-2.915] | <0.001 |
| Hemato_Disease | 221 | 8 | 3.62 | 2.87 | [1.44-5.73] | 0.002 | 1.087 | [0.522-2.264] | 0.823 |
| Neurologic_Disease | 794 | 25 | 3.15 | 2.6 | [1.73-3.90] | <0.001 | 1.1 | [0.712-1.699] | 0.668 |
| Model ROC Curve - Area 0.833 ; Asymptotic 95% Confidence Interval [0.811-0.854] |  |  |  |  |  |  |  |  |  |

**Table 3.** Association between geodemographic and pre-existing conditions and Death

| Exposure | Total | Deaths | Case fatality rate(%) | crudeRR | CI95% | P-value | aRR | [95% Conf | p-value |
| --- | --- | --- | --- | --- | --- | --- | --- | --- | --- |
| **Gender** |  |  |  |  |  |  |  |  |  |
| Female | 11900 | 253 | 2.13 |  |  |  |  |  | Ref |
| Male | 8370 | 249 | 2.97 | 1.4 | [1.18-1.66] | 0 | 1.413 | [1.189-1.68] | <0.001 |
| **Age** |  |  |  |  |  |  |  |  |  |
| 0-49 | 9675 | 4 | 0.04 |  |  |  |  |  | Ref |
| 50-59 | 3548 | 15 | 0.42 | 10.23 | [3.40-30.79] | <0.001 | 9.806 | [3.252-29.571] | <0.001 |
| 60-69 | 2459 | 44 | 1.79 | 43.28 | [15.57-120.34] | <0.001 | 37.052 | [13.273-103.432] | <0.001 |
| 70-79 | 1800 | 116 | 6.44 | 155.88 | [57.60-421.79] | <0.001 | 112.707 | [41.177-308.495] | <0.001 |
| 80-89 | 1924 | 212 | 11.02 | 266.52 | [99.23-715.80] | <0.001 | 179.145 | [65.623-489.048] | <0.001 |
| >90 | 864 | 111 | 12.85 | 310.74 | [114.88-840.52] | <0.001 | 226.802 | [82.688-622.087] | <0.001 |
| **Region** |  |  |  |  |  |  |  |  |  |
| North | 12196 | 315 | 2.58 |  |  |  |  |  | Ref |
| Acores | 48 | 0 | 0 | 0 | [..] | 0.259 | 0 | [0-0] | <0.001 |
| Alentejo | 387 | 6 | 1.55 | 0.6 | [0.27-1.34] | 0.205 | 0.8 | [0.365-1.755] | 0.578 |
| Algarve | 470 | 6 | 1.28 | 0.49 | [0.22-1.10] | 0.077 | 0.646 | [0.299-1.399] | 0.268 |
| Center | 2805 | 93 | 3.32 | 1.28 | [1.02-1.61] | 0.031 | 0.9 | [0.721-1.124] | 0.352 |
| LVT | 4261 | 74 | 1.74 | 0.67 | [0.52-0.86] | 0.002 | 0.665 | [0.525-0.842] | 0.001 |
| Madeira | 90 | 0 | 0 | 0 | [..] | 0.122 | 0 | [0-0] | <0.001 |
| **Comorbidities** |  |  |  |  |  |  |  |  |  |
| Asthma | 277 | 3 | 1.08 | 0.43 | [0.14-1.34] | 0.133 | 0.73 | [0.238-2.233] | 0.581 |
| Cancer | 603 | 47 | 7.79 | 3.37 | [2.52-4.50] | <0.001 | 1.253 | [0.921-1.704] | 0.151 |
| Cardiac_Disease | 53 | 19 | 35.85 | 15.01 | [10.36-21.74] | <0.001 | 2.576 | [1.708-3.884] | <0.001 |
| Diabetes | 1144 | 83 | 7.26 | 3.31 | [2.64-4.16] | <0.001 | 1.002 | [0.797-1.26] | 0.985 |
| Immunodeficiencies | 107 | 6 | 5.61 | 2.28 | [1.04-4.98] | 0.037 | 1.523 | [0.693-3.349] | 0.295 |
| Kidney_Disease | 400 | 98 | 24.5 | 12.05 | [9.89-14.68] | <0.001 | 2.932 | [2.329-3.691] | <0.001 |
| Liver_Disease | 107 | 7 | 6.54 | 2.66 | [1.30-5.48] | 0.007 | 0.84 | [0.36-1.961] | 0.687 |
| Lung_Disease | 686 | 60 | 8.75 | 3.88 | [2.99-5.02] | <0.001 | 1.291 | [0.987-1.689] | 0.062 |
| Hemato_Disease | 220 | 29 | 13.18 | 5.59 | [3.94-7.93] | <0.001 | 1.218 | [0.839-1.769] | 0.3 |
| Neurologic_Disease | 790 | 123 | 15.57 | 8 | [6.61-9.68] | <0.001 | 2.041 | [1.66-2.51] | <0.001 |

ROC curve -Area [0.909] ; Asymptotic 95% Confidence Interval [0.900-0.918]

### Risk Factors for ICU admission

There was a consistent increase in risk of admission to ICU with increasing age until 70–79 (aRR:10.376 CI95%:6.47–16.641), reducing in the subsequent age groups. These findings are maintained after adjustment. As for hospitalization, different regions have different ICU risks and statistically significant differences in RR that are maintained after adjustment. Among regions with cases in ICU the North Region has a lower risk compared with any other region. The diseases with higher adjusted RR for admission to ICU were Cardiac Disease (aRR:4.33 CI95%:2.53–7.41), Immunodeficiencies (aRR:2.767 CI95%:1.348–5.683), Kidney Disease (aRR:2.439 CI95%:1.61–3.697), and Lung Disease (aRR:2.035 CI95%:1.421–2.915). Liver disease and neurological disease were not associated with ICU admission although they were for hospitalization. Age groups > 60 (aRR:8.805 CI95%:5.594–13.861) had higher aRR than any chronic disease alone. The adjusted risk of admission to ICU in cases aged 70–79 was more than 10 times the risk of cases aged 0–50.

Regions maintain statistically significant differences after adjustment.

### Risk Factors for Death

Risk of death is disproportionally affected by age. We observed that there is a constant increase of risk of death with age, unlike what was seen with hospitalization and ICU admissions. Among cases aged 0–50 the case fatality rate (CFR) was 0.04%, contrasting with a CFR of 12.85% among those older than 90 years. The aRR increased more significantly from 70- 79(aRR:112.707 CI95%:41.177–308.495). Ddifferent regions had slightly different CFRs and statistically significant differences of the aRR for 3 regions. The comorbidities with higher aRR were Kidney(aRR:2.932 CI95%:2.329–3.691), Cardiac(aRR:2.576 CI95%:1.708–3.884), and Neurological disease (aRR:2.041 CI95%: 1.66–2.51).

In summary, men had significantly higher crude and adjusted RR higher than women for all outcomes. Age is the strongest risk factor after adjustment for all outcomes, especially for the outcome death. Risk of ICU and Hospital ward admission increased with age, but was reduced in most advanced ages groups. Comorbidities all have similar risk for hospitalization but risk increases for some of them and decreases for other for ICU and death as outcomes. Regions maintain small but significant differences for all outcomes after adjustment but with weaker associations for death.

## Discussion

This study was a first attempt at identifying demographic and clinical factors associated with adverse outcomes of COVID-19 in the Portuguese population: hospitalization, admission to ICU and death. It has the benefits of presenting Relative Risks adjusted for confounding, and uses good quality data from all cases in the country and not only cases in specific clinical settings such as hospitals.

We used information extracted from the National Epidemiological Surveillance System (SINAVE) electronic platform, which is thoroughly validated by the Public Health authorities network in Portugal. This ensures a good level of data accuracy, data completeness and representativeness, allowing generalization of findings to the general population. Still we most consider surveillance system sensitivity could be lower before March 26 as the testing strategy was less broad. Some regional differences in testing strategies probably remained even though we accounted for this possible confounding.

In this study we found that increasing age is, as in other multivariable analysis,^9 11^ the most relevant risk factor for Hospitalization, ICU admission and death^11^ and is disproportionately high for the outcomes death comparing to comorbidities alone with a fast increase after 70. The very high values of aRR for older ages in comparison with other risk factors can be justified by the very low fatality observed in the reference age group 0–50.

Hospitalization and ICU admissions had a relevant increase in risk in 60–69 and 70–79. The risk of ICU admission reduces after 70–79. This is not expected to be due to any negative selection based on age since ICU beds occupation did not go over 60%^19^ during that period and there are guidelines with criteria for ICU admission issued by DGS^20^ that consider only clinical severity and no age criteria. Other studies found similar situations for influenza^21 22^. It is possible that some older patients may die without meeting criteria for ICU admission or eventually, many who end up meeting those criteria may die before they can be admitted. However further research is needed to understand the benefit of intensive care among the very elderly. Debate has been ongoing on this topic considering the challenges and ethics of admitting the very elderly to ICU, patient and family wishes and therapeutic futility^23 24 25 26 27^.

Older age was by far the most important determinant for Covid-19 associated death.

Most comorbidities we investigated were associated with increased risk for hospitalization, admission to ICU and death - especially cardiovascular, kidney, respiratory and neurologic disease - but they vary for different outcomes. All comorbidities increased risk more homogenously for hospitalization than for ICU and Death. This could be explained by lower thresholds for the decision to admit patients to a general ward vs ICU and by Portuguese guidelines that consider the existence of comorbidities for hospital admission but included mainly clinical severity criteria for ICU^20^. Risks factors for death vary from other outcomes partially because death does not include a clinical management decision. For ICU admission, most relevant risk factors besides age were Cardiac disease, Immunodeficiencies, Kidney Disease, and Lung disease. Liver, Neurologic and Hematologic Disease were not significantly associated).

Asthma was not a risk factor for any outcome, in line with what was found in other studies^9 10^. However he largest cohort investigated to date found a slight increase in adjusted risk of death for asthma, especially if severe^11^.

We found particularly strong associations of older age, cardiac disease and chronic kidney disease at risk of both ICU admission and death, curiously with slightly less influence from chronic lung disease as in a similar study^9^ even though the largest cohort study for outcome death found that lung disease had the highest risk among comorbidities^11^.

For the outcome death we found similar results to the ISARIC study^10^ and to the OpenSafely Project Cohort^11^ although the differences in risk measures for age are more expressive.

The comorbidities with higher risk vary for different outcomes. Immunodeficiencies, Cardiac, Kidney, and Lung Disease gain importance for the outcome ICU comparing to their risks for hospitalization. For death the strongest risk factors were Kidney, Cardiac and Chronic Neurological Disease.

Regions were included in the models primarily to minimize unobserved confounding and we found small differences in Region’s association with outcomes after adjustment for age and comorbidities. Since some heterogeneity naturally exist between Regions epidemic situations, testing strategies and case-ascertainment, we hypothesized that there could be unobserved confounding mediated through regions^9^. Some small regional differences remain after adjusting for other covariables, and are more relevant for the outcomes Hospitalization and ICU even if they exist for death. This finding may be due non observed confounding factors. It is probably primarily related to different regional detection rates among mild and asymptomatic infection in different epidemic stages. Significant differences in healthcare services response are not expected as Portugal has a good hospital network, no region surpassed hospital capacity and guidelines with criteria for admission to general ward and Intensive Care have been issued by the Directorate-General of Health^20^. In the outcome data, it is possible that differential delay in registry of outcomes in SINAVEmed database from different regions may also have contributed to small regional differences eventually more importantly for death, because of the larger delay from initial notification to outcome, although we believe that eventual bias would be small.

Interestingly, ICU criteria are based on clinical severity indicators while for admission in a general ward the existence of comorbidities alone is a criteria for admission (namely StageV Kidney Disease, Immunodeficiencies, active malignancy, and decompensated Chronic Lung Disease, Asthma, Heart Failure, Diabetes e Cirrhosis(Liver Disease). This may partly explain why some of this comorbidities are risk factors for hospital admission in general ward but not for ICU or death.

Number of tests in asymptomatic patients in specific settings such as nursing homes and healthcare workers may have varied across regions. Since under-ascertainment can have an important impact on the proportion of outcomes by strata, and considering regional and age specific differences in case ascertainment, healthcare services practices and availability in different epidemic phases, we believe the adjustment for Regions may be relevant for this type of analysis but differences in regional risk should be interpreted with caution, specially with small number of cases and small risk differences.

The study has some limitations. The extracted data is related to an early phase of the epidemic in Portugal and as such, risks associated may still change with increased testing and detection of mild and asymptomatic infection, changes in Regional epidemic behaviour and further data validation. There are relevant comorbidities that we could not adjust for that have been previously found to be of relevance for the COVID-19 severity outcomes, such as obesity^9 10 11^, economic deprivation and black and minority ethnic groups. Hypertension is also not included since it was not available in SINAVEmed dataset, although recent evidence form the largest cohort to date suggest that controlled Hypertension (HT) alone does not increase the risk of death of COVID-19 patients^11^. A study that found increased risk for Hypertension did not adjust for age, which we know is strongly associated with risk of death and severe disease^29^. In our study no data on smoking was available but, as with hypertension, it does not seem to be a relevant factor for poorer outcomes in the OpenSafely Project Cohort, where some evidence of increased risks in former smokers was found but no effect for actual smokers.^11^

Cardiovascular disease small numbers are probably related to the fact that, contrary to other Comorbidities, Cardiac Disease is not in a specific field in the SINAVEmed notification form. As such physicians would need to describe that condition on the field “others chronic conditions”. This may introduce information bias as the field “other chronic conditions” may be more frequently completed for cases with poorer outcomes, overestimating the risk factor Cardiac Disease. However, other studies conducting multivariable analysis from cases have found Cardiac Disease to be one of the most relevant risk factors.^9 10 11^

Cases were not censored considering time of follow-up and final outcomes. As such it is possible that some cases in the dataset could still developed one of the analysed outcome. We believe this would not introduce a systematic error in risk estimates for age and comorbidities.

We used adjusted relative risks in multivariate analysis using Poisson regressions assuming constant time of exposure. The use of an adjusted odds ratio to estimate an adjusted relative risk can be appropriate for studies of rare outcomes(< 10%) but may overestimate the risk if outcomes are more frequent and can be more valid then the odds ratio to represent a risk ratio especially if we want to estimate the effect of specific exposures and could overestimate the risk.^30 31 32^. Overestimating RR could inappropriately affect clinical decision-making,policy development, and errors in economic evaluation of potential intervention and prevention programs or treatments.^30 31^ Poisson regression is likely to compute confidence intervals that are conservative with more common outcomes, suggesting less precision than is true.

Age was categorized in 10 years after 0–49 age groups for all the outcomes. Other studies conducting multivariable analysis have used such age categorization to identify risk factors for COVID-19 clinical outcomes “including the largest cohort studied for risk factors for death from COVID-19^11^. Age 0–50 was used as reference category as in previous studies very low risk was found in this group for severe outcomes.^9 10 11^

Our findings may help shaping stratified public health policy, modelling risk criteria for clinical management and aid in definition of epidemic scenarios for the assessment of health needs in the face of different possible evolutions of the epidemic.

Clinical risk assessment tools may be built to aid clinical decisions related to admission to hospital or to ICU, since most patients can be safely followed up at home by healthcare professionals, by phone. Most EU countries took this approach.^33^

Policy and recommendations in Portugal and other countries consider specific comorbidities and age cut-offs to consider people at risk. As such it is relevant to understand what is the level of risk added by each specific characteristic, accounting for differences in case detection in different age groups and healthcare services heterogeneity to shape policy and define risk categories for health-care practitioners, organisations, and the general public.

In Portugal patients with Hypertension, diabetes, cardiovascular disease, chronic respiratory diseases, oncological diseases and patients with renal failure, may have medically justified absences from work if they cannot work from home^34^. We could show that if these patients become ill with COVID-19 they will be at increased risk of having a poor outcome except for hypertension, but would probably not be found to be a risk factor after adjustment considering recent evidence^11 13^. However no age categories are referred in this legal document, (even tough age risks are referred in other guidance and recommendations) and higher age groups pose a higher risk than comorbidities for all outcomes starting from 60 but specially after 70 for all considered outcomes but higher for death.

In terms of risk estimates for absolute risks of specific outcomes among infected, including for individual risk estimates using generated model parameters but also for model inputs to estimate healthcare demand scenarios we should consider under-ascertainment of mildly symptomatic and asymptomatic cases.^35^ Furthermore, under-ascertainment is probably higher in younger ages and rises with age as they may have a lower threshold for testing and have a smaller proportion of mild and asymptomatic infections. This could mean modelled absolute risk estimates are overestimated in all age groups but specially in the younger population.

It will be relevant to revaluate the same data later in time to see if the findings are maintained and what changes may happen.

## Conclusions

Advancing age was the most relevant risk factor for all outcomes and grows faster after 60 for the outcome death, as risk is low in those aged 0–50.Control strategies should be based on an attempt to reduce the number of serious cases and deaths. For this, it will be important to reinforce measures that prevent infection in people over 60 but mostly over 70 years of age. A recent study suggest that lockdown policies targeted at sheltering and testing the elderly have huge gains in lives saved and hospitalizations at low economic cost and large returns to broad testing policy.^36^ Good epidemiological surveillance of settings with higher risk population, and long-term care facilities is part of an efficient way to achieve this goal and has been promoted in recent months.

Comorbidities also have an impact on clinical outcomes (especially Cardiac, Kidney, Lung disease,Immunodeficiencies and Neurologic Disease) but they are smaller than age and vary for different outcomes.

Risk stratified public health measures should consider age primarily although individual preventive behaviours should be promoted across all age groups to reduce overall spread and ultimately prevent infection in higher risk population.

## Data Availability

Anonymized Data was shared by Directorate-General of Health with the NAtional School of Public Health

## Acknowledgements

Paula Vasconcelos, Vera Pereira Machado, Pedro de Vasconcelos Monteiro for relevant inputs.

